# Factors Associated with Medicine Timing Effects: A Meta-analysis

**DOI:** 10.1101/2021.10.24.21265348

**Authors:** Marc D. Ruben, Lauren J. Francey, Gang Wu, David F. Smith, Garret A. FitzGerald, Jacob J. Hughey, John B. Hogenesch

**Affiliations:** Divisions of Human Genetics and Immunobiology, Department of Pediatrics, Cincinnati Children’s Hospital Medical Center, Cincinnati, USA; Divisions of Pediatric Otolaryngology and Pulmonary and Sleep Medicine, Cincinnati Children’s Hospital Medical Center, Cincinnati, USA; Department of Otolaryngology-Head and Neck Surgery, University of Cincinnati School of Medicine, Cincinnati, USA; Department of Systems Pharmacology and Translational Therapeutics, at the University of Pennsylvania Perelman School of Medicine, Philadelphia, USA; Department of Medicine, at the University of Pennsylvania Perelman School of Medicine, Philadelphia, USA; Institute for Translational Medicine and Therapeutics (ITMAT), at the University of Pennsylvania Perelman School of Medicine, Philadelphia, USA; Department of Biomedical Informatics, Vanderbilt University Medical Center, Nashville, USA; Department of Biological Sciences, Vanderbilt University Medical Center, Nashville, USA

## Abstract

**Importance:** Clinical evidence suggests that the time of day of treatment can affect outcomes in many different diseases, but this information is dispersed, imprecise, and heterogeneous. Consequently, practice guidelines and clinical care recommendations seldom specify intervention time.

**Objective:** To understand the sources of variability and summarize clinical findings on the time of day effects of medicine.

**Data Sources:** A systematic search of Pubmed, Google Scholar, and ClinicalTrials.gov for “chronotherapy” OR “time of administration”.

**Study Selection:** Any clinical study since 2000, randomized or observational, that compared the effects of treatment at different times of day. We included pharmacologic or surgical interventions having at least one continuous outcome.

**Data Extraction and Synthesis:** For selected studies, we extracted the mean and variance of each time-of-day treatment group. From these, we computed the standardized mean difference (SMD) as the measure of timing effect. Where a study reported multiple outcomes, we selected a single outcome based on a defined order of priority.

**Main Outcomes and Measures:** We estimated overall pooled effect size and heterogeneity by a random effects model, followed by outlier detection and subgroup analyses to evaluate how study factors, including drug, design, outcome, and source, associate with timing effect.

**Results:** 78 studies met the inclusion criteria, comprising 48 distinct interventions over many therapeutic areas. We found an overall effect of time on clinical outcomes but with substantial heterogeneity between studies. Predicted effects range from none to large depending on the study context. Study size, registration status, and source are associated with the magnitude of effect. Larger trials and those that were pre-registered have markedly smaller effects, suggesting that the published record overstates the effects of the timing of medicine on clinical outcomes. In particular, the notion that antihypertensives are more effective if taken at bedtime draws disproportionately from one source in the field, which consistently detects larger effects than the community average. Lastly, among the most highly studied drug timing relationships, aspirin’s anti-clotting effect stands out, consistently favoring evening over morning dosing.

**Conclusions and Relevance:** While accounts of drug timing effects have focused on *yes/no*, appreciating the range of probable effects may help clarify where ‘circadian medicine’ meets the threshold for clinical benefit.

## Introduction

Half of human genes and the physiology they control are expressed with a 24 h rhythm with implications for medicine ^1,2^. Treatment time effects can in theory arise from circadian influences on pharmacokinetics, pharmacodynamics, or underlying physiology. Indeed, dozens of clinical trials suggest that time of dosing can affect responses in cancer, heart, lung, and other diseases. Even vaccines may be influenced by the timing of their administration ^3,4^. Yet, outside of sleep medicine, time of day is rarely considered in medical practice. The most recent WHO Model List of Essential Medicines makes no mention of dosing time. In view of notable advances in circadian biology research, why are there so few examples of clinical use?

Dosing time has been most studied in hypertension, and randomized trials seem to converge on a few fundamentals ^5–7^. For example, multiple randomized trials show that short-acting antihypertensives taken at bedtime better restore the normal overnight drop in blood pressure compared to morning dosing ^6^. But even here there is uncertainty. Trials differ by drug, design, and endpoint, and not all findings point in the same direction. Of the dozens of antihypertensive medications, none have garnered an FDA time-of-day directive, and the importance of dosing time is disputed ^8–12^. In fact, we have only a superficial understanding of how much treatment time affects cardiac outcomes; this question applies to all indications. A history of trials report treatment time differences, but information is heterogeneous and imprecise. There is a need to synthesize the evidence.

To understand the sources of variability and more adequately summarize clinical findings on the time-of-day uses of medicine, we analyzed 78 studies that compared treatment at different times of day. This comprised 48 distinct interventions over many therapeutic areas. To our knowledge, this is the largest such analysis to date.

## Methods

### Study Identification

We conducted a systematic search of PubMed, ClinicalTrials.gov, and Google Scholar for clinical studies that compared the effects of treatment administered at different times of day in accordance with the Preferred Reporting Items for Systematic Reviews and Meta-analyses (PRISMA) reporting guideline. The first search was performed in March 2020. We used the search terms “chronotherapy” or “time of administration” and included only articles in English.

After title and abstract screening, the full texts of potentially relevant studies were reviewed and selected if they evaluated the effects of the same treatment given at different times of day. We allowed any pharmacologic or surgical intervention as long as the study tested at least one continuous outcome measure. Because the aim of our study was a broad evaluation of medicine timing effects, we allowed any study design––randomized or observational, and any disease context, study population, and outcome/endpoint––as long as the effect mean and variance of each time-of-day group were available. The only exception to this were studies addressing sleep disorders, which we excluded.

We screened the hits from the initial search for relevant references. We performed a final search of the same three databases in June 2021 (Fig. 1 Supplement). Our final selection includes studies meeting the above criteria published since January 1st 2000.

### Data Extraction and Methodological Quality Assessment

This study was not submitted for institutional review board approval because it did not involve individual patient information and all data extraction was from publicly available sources. We collected study characteristics: drug ingredient, drug class, drug half-life, study design, registration status, treatment setting, patient demographics, investigator source, and primary and secondary outcomes. For study outcomes, we extracted the size, mean, and variance of each time-of-day treatment group.

While we did not grade the quality of each study a priori, we collected study characteristics used by established methods of grading ^13^. We sought to understand sources of heterogeneity in the body of literature and our analyses consider several commonly used proxies for quality.

### Statistical Analysis

We computed Cohen’s d standardized mean difference (SMD) for each continuous outcome, from each study ^14^. Specifically, we subtracted the mean post-evening-treatment outcome from the mean post-morning-treatment outcome and divided the result by the pooled standard deviation for both groups (Eq. 1 and Eq. 2). Where more than two times of day were tested, we computed the SMD between the two groups with the largest raw mean difference.

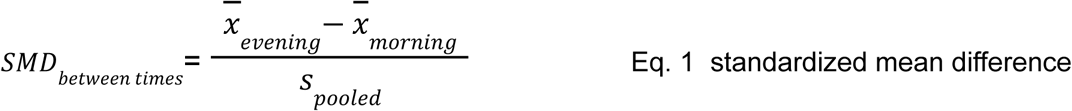

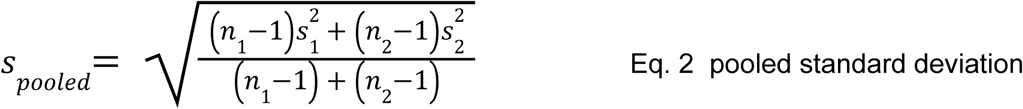

n_1_, s_1_: sample size and standard deviation of evening group

n_2_, s_2_: sample size and standard deviation of morning group

To correct for small-sample bias in Cohen’s SMD, we converted Cohen’s SMD to Hedge’s SMD ^15^ using the R package ‘esc’ ^16^. Standard error (SEM) of SMD was computed according to Eq. 3 ^17^. Datafile 1 in the supplement contains the fully curated and coded dataset.

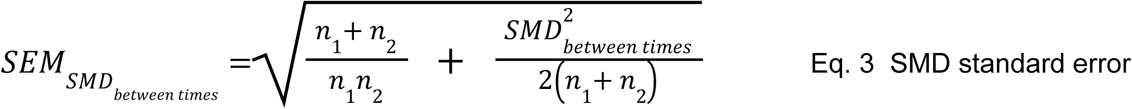

We prioritized end-of-study means as opposed to baseline subtracted, ie. change score, means. In rare cases where means were baseline-covariate-adjusted, we prioritized these, as reported in Data file 1 in the supplement. In cases where within-group variances were expressed as standard error of the mean (SEM) or 95% confidence intervals, we computed SD according to methods described in the Cochrane Handbook for Systematic Reviews of Interventions ^18^. If only median and interquartile range were reported, mean and SD were approximated ^19,20^.

Where a study reported multiple continuous outcomes, we selected a single outcome based on the following order of priority: (i) significant effect (p < 0.05) *and* a designated primary endpoint, (ii) designated primary endpoint, (iii) significant effect, and lastly (iv) none of the above. If multiple outcomes remained, we selected the one with the largest SMD to include in the meta-analysis.

In a small number of cases, we excluded certain outcomes from eligibility (Data file 1 in the supplement). For example, whereas awake/asleep ambulatory blood pressure are well accepted clinical endpoints in a study of hypertension, mean blood pressure over an arbitrarily defined window, eg. 11AM to 3PM, is not.

In our primary analysis, we estimated the overall SMD across studies by pooling the individual study SMDs using a random-effects model with a DerSimonian-Laird estimator ^21^ via the R package ‘meta’ ^22^. To identify potential sources of heterogeneity between studies, we recalculated the meta-analysis results N times, each time leaving out one study, and then prespecified subgroup analyses were performed using a mixed-effects model via the R package ‘dmetar’ ^23^. The mixed-effects model estimates the variability within subgroups as a random effect and the variability between subgroups as a fixed effect. A p-value for difference between groups was calculated via Q-test. Subgroups evaluated include study design, registration status, source, and drug kinetics. Drug half-lives were extracted from DrugBank 5.0 ^24^, FDA-label, or research publication. We evaluated for small-study bias via funnel plot ^25,26^.

Our study includes two secondary analyses. The first compares *raw* mean differences among a set of blood pressure studies with outcomes on the same unit scale, by drug type and kinetics, patient demographics, and the study source. The second analysis evaluated the reproducibility of interventions tested in more than one study.

## Results

Our search revealed 172 potentially relevant reports, of which 78 fulfilled our eligibility criteria (Fig. 1 Supplement). The 78 studies encompass 48 distinct treatments tested at different times of day in multiple disease settings (Figure 1A, Data file 1 Supplement, Methods). Most studies tested only two time points, typically morning and evening.

**Fig. 1.**
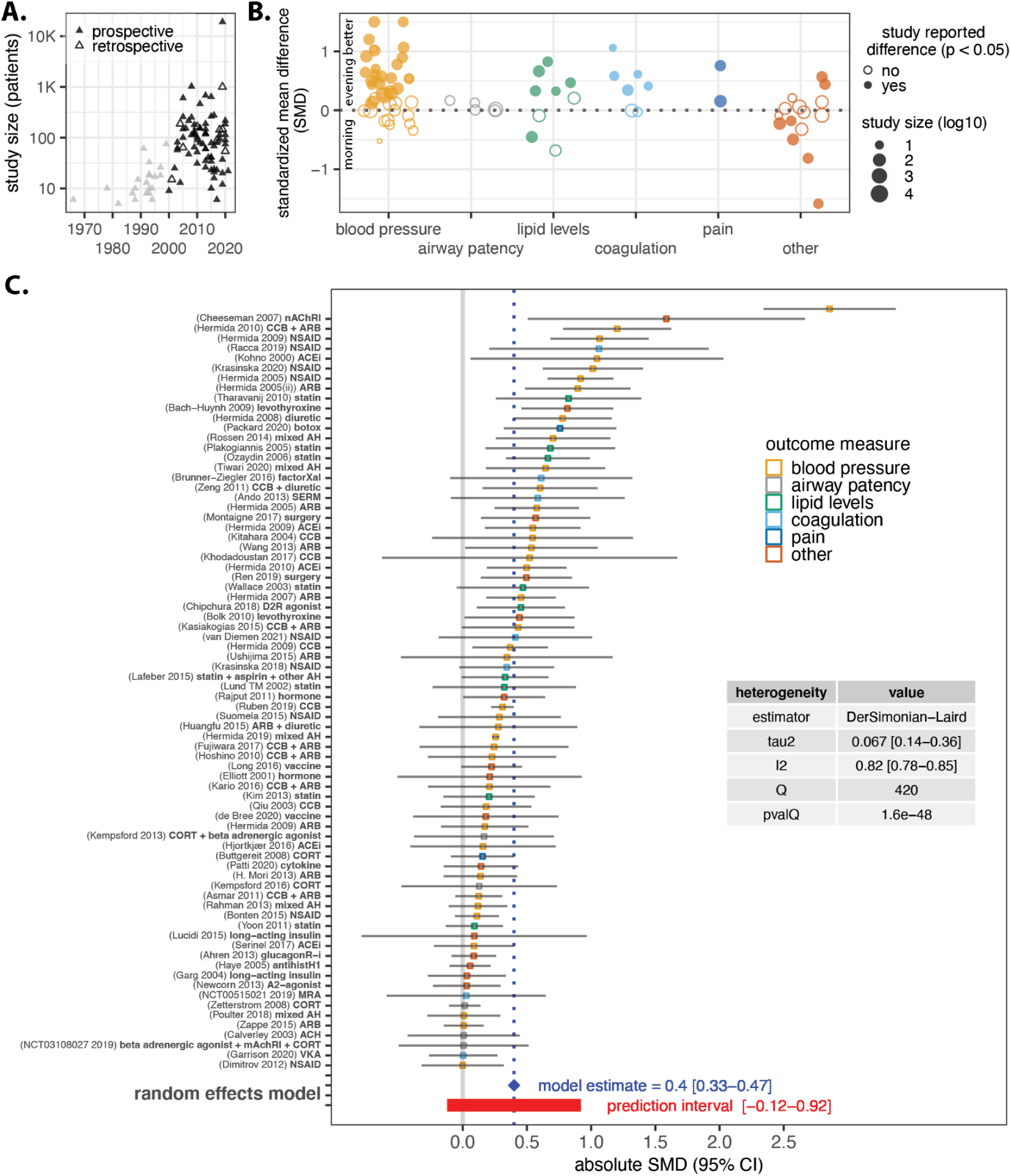
Effects of intervention time in clinical studies. (**A**) 78 studies since 2000 (black triangles) that tested the impact of time of day on treatment outcomes. Data file 1 in the Supplement holds the full curated dataset. (**B**) Standardized mean difference (SMD) grouped by the type of outcome measure, where 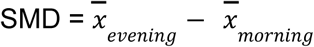 divided by the pooled standard deviation. *Filled circles*: outcomes that were reported in the original research as significant at p < 0.05 by null hypothesis testing. SMD values were capped at 1.5 for visualization, impacting two blood pressure studies with SMDs of 6.7 and 2.9, both set to 1.5 in this plot. All raw data is available in Data file 1 in the Supplement. (**C**) Absolute SMD with 95% confidence interval. *Blue diamond*: pooled effect under a random effects model. *Red bar*: Prediction interval within which future studies can be expected to fall. *Grey vertical line:* zero timing effect. *Inset table*: between-study heterogeneity. Note: We removed one extreme outlier study for visualization purposes only, Farah et al 2013, with SMD = ∼7.

### Standardized effects of time across interventions

We hoped to gain a sense of which interventions were most susceptible to time of day effects. We computed the standardized mean difference (SMD) ^14,17^ for each study (Fig. 1B). SMD provides a measure of effect size that is relative to variance within the study. Specifically, we subtract the mean of one time-of-day group from the other and divide the result by the pooled standard deviation (SD) (Methods). Effect sizes are given as a fraction of the standard deviation. This measure is independent of the units or scale in the original study and allows for comparisons across studies. A study with SMD = 0.5 indicates the two groups differ by 0.5 SD, SMD = 1 indicates the two groups differ by 1 SD, and so on. Where a study reported multiple outcomes, we selected a single outcome based on an order of priority described in Methods.

SMDs range from −0.16 to 6.7 (Fig. 1B), with positive values indicating that evening dosing was favorable to morning dosing, and vice versa. Evening dosing was favorable in most studies (68%). SMD alone, however, does not indicate sampling error, as studies differ in precision. To estimate the overall effect of time, we pooled all studies together under a random effects model which weights studies based on their estimated precision (Fig. 1C). The pooled estimate is therefore a measure of the mean of a distribution of true effects ^17,27^. The estimated absolute SMD of 0.41 (95%CI: 0.34 - 0.49, p < 0.0001) indicates an overall positive effect of time. However, model indicators ^28^ suggest substantial between-study heterogeneity (Fig. 1C; I^2^ = 82%, ie., the percent of variability not caused by sampling error). The wide prediction interval ^29^ from −0.12 to 0.94 means that the pooled estimate of 0.41 is unlikely to be robust in every context. We expect some future studies will find little or no timing effect, whereas others will find large effects. This led us to ask which studies were more or less influential in this prediction.

### Leave-one-out analysis identifies an investigator source of heterogeneity

To identify sources of heterogeneity between studies we recalculated the meta-analysis results N times, each time leaving out one study. This allows us to detect the contribution of each study to the overall pooled effect (Fig. 2A, left). Specifically, the ‘difference in fit(s)’ (DFFITS) metric ^30^ quantifies how much and in which direction the pooled effect changes when a study is omitted. Among the 12 most effect-lowering studies, nine of them relate to blood pressure, seven of which draw from sites in Northwest Spain (N.W. Spain) involving the same investigator (Fig. 2A, middle). This source is responsible for 13 of the 39 blood pressure studies in this meta-analysis, with the other 26 studies spread across different investigative groups and locations (Data file 1 Supplement). Using a mixed-model approach that pools the effect of subgroups and compares between them ^31^, the pooled effect from N.W. Spain is 0.80 (95%CI: 0.52 - 1.1) compared to 0.33 (95%CI: 0.24 - 0.41) for all others (p < 0.005) (Fig. 2B). We explore this further in a later section.

**Fig. 2.**
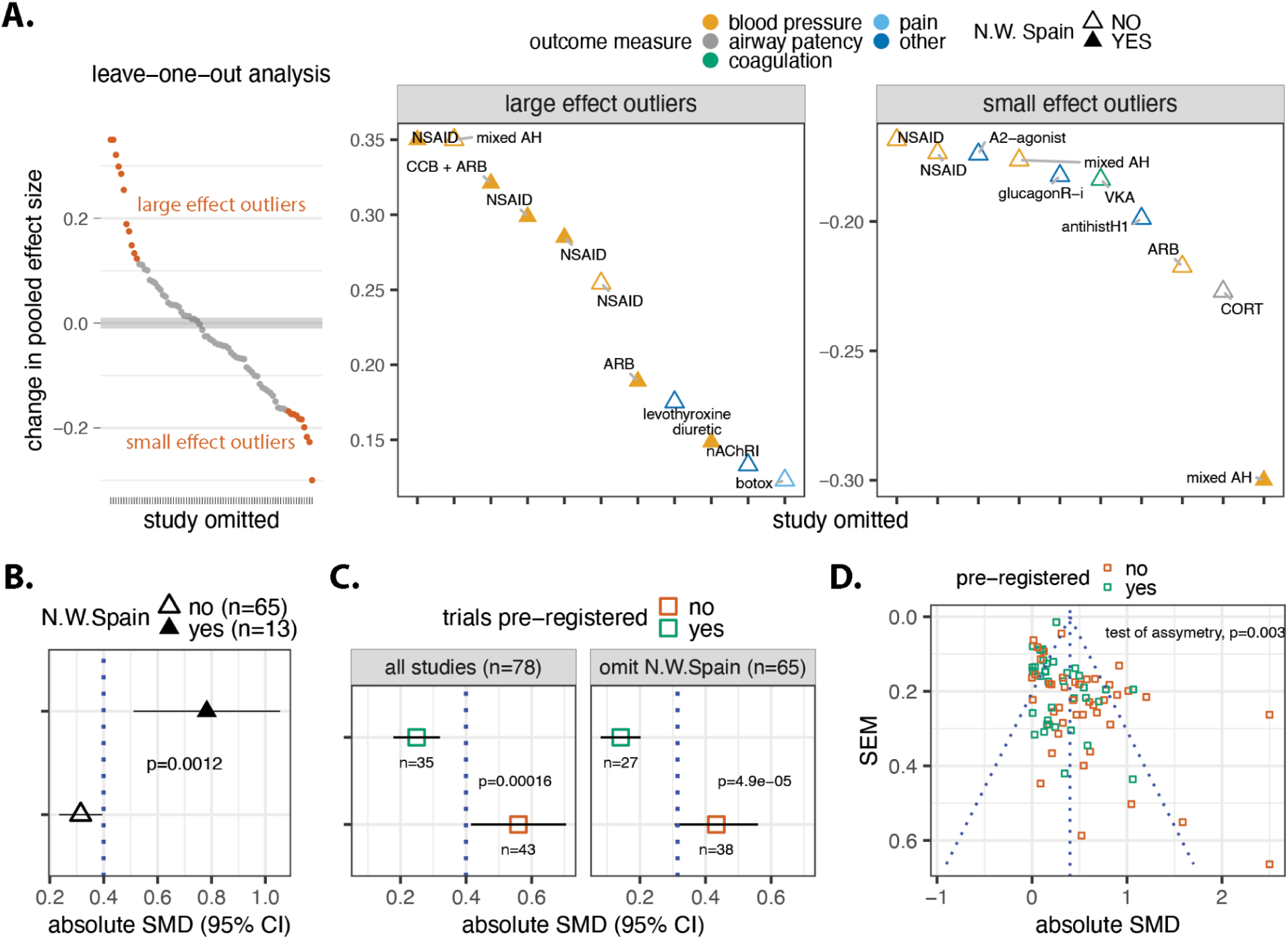
Study source and pre-registration condition effect size. (**A**) Size and direction of the change in the pooled effect when each study is omitted. DFFITS on the y-axis is expressed in standard deviations; larger values indicate decreased pooled effect when the study is omitted, and vice versa. Top 10 increases and decreases labeled. DFFITS values were capped at 0.35 for visualization, impacting two blood pressure studies with values of 6.7 and 2.9. (**B**) Subgroup analysis of Northwest Spain (N.W. Spain) vs.other studies. *Blue line:* pooled effect of all studies. (**C**) Subgroup analysis of pre-registered versus non-pre-registered for all studies (left panel) or omitting N.W. Spain (right panel). *Blue line:* pooled effect for all studies. (**D**) Study precision (SEM_SMD_) versus effect (SMD). *Blue vertical line:* pooled effect. *Blue diagonal lines:* pseudo 95^th^ percentile confidence intervals around the pooled average as precision decreases from zero. P value from Egger’s test for asymmetry of the funnel plot. Absolute SMD values were capped at 2.5 for visualization only, impacting two blood pressure studies, SMD = 6.7 and 2.9, both set to 2.5 in this plot. *Drug class abbreviations:* ARB: angiotensin receptor blocker, CCB: Ca^2+^ channel blocker, nAChRi: anticholinergic, NSAID: non-steroidal anti-inflammatory, mixed AH: combination of antihypertensives, ACEi: angiotensin converting enzyme inhibitor; A2-agonist: alpha-2 adrenergic receptor agonist, CORT: corticosteroid, antihistH1: antihistamine, GlucagonR-i: glucagon-like peptide-1 receptor agonist, VKA: vitamin K antagonist.

### Timing effects are associated with registration status

We evaluated whether there are other study contexts where effects are lower or higher. Using a mixed model approach as above, we tested the impact of study pre-registration and design.

Clinical study registration provides transparency and discourages publication bias. Because only 34 of 78 studies in this analysis were prospectively registered on ClinicalTrials.gov or an international equivalent, we speculated that the overall evidence base may be distorted. The idea is that without a public record, negative or null trial results may be less likely to be published ^32,33^. Indeed, the pooled effect of pre-registered studies is 0.26 (95%CI: 0.19 - 0.34) compared to 0.57 (95%CI: 0.43 - 0.72) for non-registered studies (p < 0.001) (Fig. 2C). The pooled effect of pre-registered studies becomes even smaller (0.16, 95%CI: 0.09 - 0.22) if we omit all studies from N.W. Spain, several of which were pre-registered.

We found no evidence of association between study design and effect (Fig. 1A Supplement). Although parallel trials had a larger effect than crossover or retrospective designs, this difference can be explained by N.W. Spain studies, most of which were parallel designs.

### A small-study effect may indicate publication bias

There is a tendency for smaller studies in a meta-analysis to show larger effects ^34,35^. One possible cause is reporting bias, whereby small negative studies are less likely to be published. Funnel plots are commonly used to investigate small-study bias. The principle is that as studies increase in size and precision, their location around the pooled average effect narrows, forming a funnel. In the absence of bias, small (less precise) study effects should scatter widely, with spread narrowing for larger (more precise) studies. Our analysis detects asymmetry indicative of small-study bias (Fig. 2D). Specifically, the largest studies fall to the left of the pooled effect. This imbalance, together with the prior observation that pre-registered studies have smaller effect sizes, suggests that the published record overstates medicine timing effects.

### The evidence for medicine timing in hypertension

Half of the studies in this meta-analysis concern the timing of antihypertensives. More than one-third of these antihypertensive studies originate from N.W. Spain. Understanding *why* findings from N.W. Spain appear as outliers is important from both a research and clinical perspective.

We first evaluated whether N.W. Spain’s large effects may be due to lower study variances. SMD is scale-free and not influenced by the units of measurement. However, choices made by an investigator, such as enrolling a narrow patient population, can reduce study variance, which may cause SMD to appear larger ^36,37^. Therefore, we computed raw mean differences (RMD = mean mmHg_morning_ – mean mmHg_evening_) for each of the 38 studies with blood pressure outcomes on the same scale. We focused on nighttime blood pressure because of its prognostic ^38–40^ and circadian ^41–44^ relevance. Overall, RMDs suggest that evening dosing is preferable to morning dosing for lowering nighttime blood pressure (Fig. 3A). However, the median RMD from N.W. Spain (5.5 mmHg) is markedly larger than other studies (1.7 mmHg). Put another way, N.W. Spain is also an outlier by RMD, as it was by SMD. Perhaps interestingly, N.W. Spain studies do have lower variances, ie. pooled standard deviations (Methods) (Fig. 3A) on average, but this is unlikely to explain the large discrepancy in RMD.

**Fig. 3.**
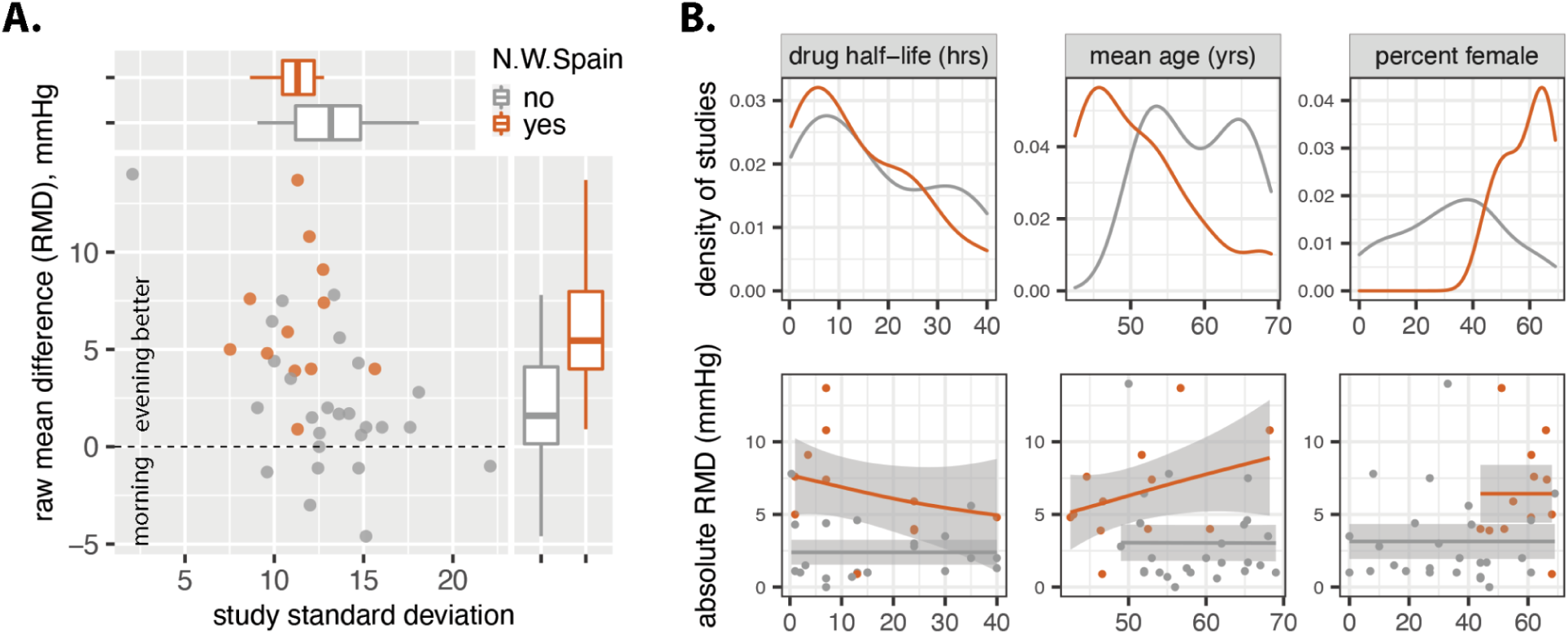
Factors influencing dosing time effects on nighttime blood pressure. (**A**) 38 studies that tested the effect of morning versus evening dosing on nighttime systolic blood pressure. Effect is shown on the vertical axis as raw mean difference (RMD) where RMD = mean mmHg_morning_ – mean mmHg_evening_. Study variance is shown on the horizontal axis as pooled standard deviation (Methods). Marginal boxplots summarise how RMD and variance are distributed. (**B**) *Top row*: density plots indicate relative numbers of studies at different values of drug half life, mean patient age, and percent sex. *Orange lines*: N.W. Spain; *grey lines*: all other studies. *Bottom row*: regressing RMD onto drug half-life, mean patient age, and percent sex. Smooth lines fit by the generalized additive model y ∼ s(x, bs = “cs”, knots = 5), where y = RMD, x = covariate, cs = cubic spline basis function. Grey shading: 95% confidence intervals. Drug half-lives obtained from DrugBank 5.0, FDA-label, or research publication. For studies with drug combinations, the shortest-acting ingredient is considered.

Can differences in the types of drugs studied explain the discrepancy? N.W. Spain has larger RMDs for all six mono- or dual-therapies that were also studied by at least one outside group (Fig. 1B Supplement). Further, the distributions of study drug half-lives are virtually indistinguishable between N.W. Spain and others (Fig. 3B, top left). Therefore, differences in the nature of the drugs studied cannot explain the discrepancy.

We next evaluated patient composition. N.W. Spain consistently enrolled a larger share of younger and female patients than other studies (Fig. 3B, top middle and right panels). This difference in study makeup is dramatic, and sex-specific effects of drug timing have been observed in other contexts ^45,46^. However, closer inspection of the data refutes this idea, as neither sex nor age track with RMD (Fig. 3A, bottom middle and right panels). In sum, the idea that antihypertensives, even short-acting, are more effective if taken at night is only weakly supported by the broader research community.

### Aspirin’s time-dependent anti-coagulative effect stands out

This meta-analysis includes studies in many conditions other than hypertension. We evaluated if there are particular areas of medicine with orthogonal lines of support for the importance of time. There were too few studies per outcome to justify regression-based subgroup analyses, so we simply visualized effect (SMD) versus precision (SEM) for each of the 38 non-hypertension studies. With the exception of airway patency, where studies can be characterized as high precision but small effect, there are no remarkable differences between outcomes (Fig. 4A, left).

**Fig. 4.**
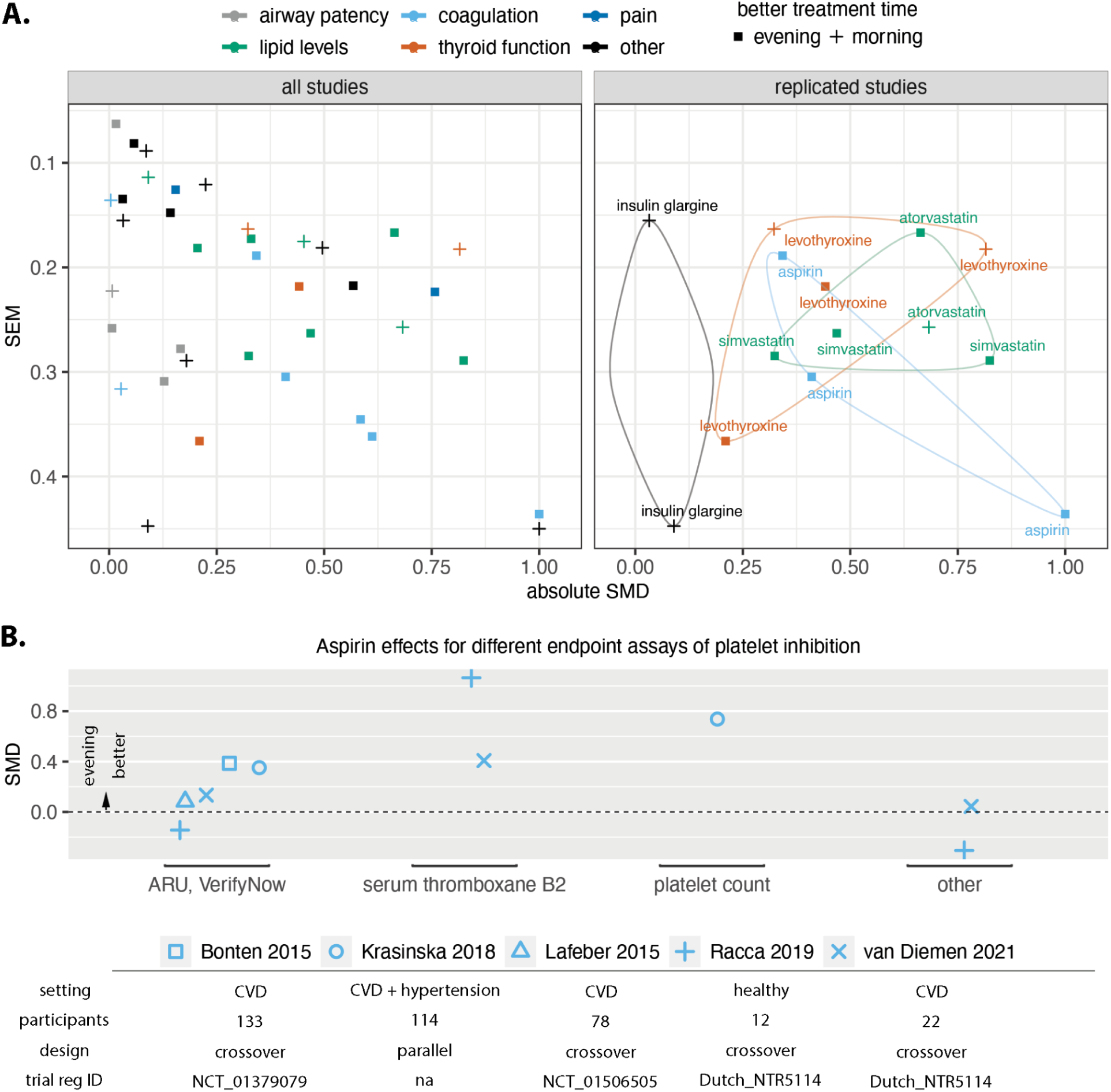
Assessment of reproducibility. (**A**) *Left*: absolute SMD versus precision (SEM) for all meta-analysis studies, excluding blood pressure outcomes (n = 38). *Point shape*: whether morning or evening treatment was more effective. *Right*: the five intervention::outcome pairs tested in more than one study: (i) aspirin::coagulation, (ii) levothyroxine::thyroid function, (iii) insulin glargine::glucose levels, (iv) simvastatin::lipid levels, and (v) atorvastatin::lipid levels. Pairs are encircled to highlight group coherence. (**B**) All outcomes of aspirin in coagulation for each of the five studies of aspirin in this meta-analysis (n = 10). Data presented here deviate from our meta-analysis inclusion criteria of one outcome per study (described in text, Methods). Point *shape*: study reference. *Abbreviations:* ARU: aspirin relative units from VerifyNow test.

We next assessed reproducibility, focusing on the five intervention::outcome pairs tested in more than one study: (i) aspirin::coagulation, (ii) levothyroxine::thyroid function, (iii) insulin glargine::glucose levels, (iv) simvastatin::lipid levels, and (v) atorvastatin::lipid levels (Fig. 4A, right). Simvastatin and aspirin were consistently more effective when taken in the evening, whereas insulin glargine was more effective when taken in the morning. Levothyroxine and atorvastatin were inconsistent, favoring evening dosing in some studies and morning in others.

All aspirin studies in this meta-analysis tested low 75-100 mg ‘cardiopreventive’ doses. Aspirin has well established anti-thrombotic efficacy and low-dose aspirin is standard-of-care for secondary prevention of cardiovascular disease ^47^, taken daily by millions of patients. The understanding that its effect is sensitive to timing is not reflected in current dosing guidelines. Given the potential clinical importance, we examined all available evidence. As a reminder, we selected only a single outcome per study to include in the meta-analysis, based on a defined order priority (Methods). However, secondary and other endpoints in a study are also informative. To scrutinize all available evidence, we compared all of the reported outcomes of aspirin in coagulation for all aspirin studies in this meta-analysis (Fig. 4B). Eight out of ten outcomes favored evening over morning dosing, across different assays of platelet inhibition.

Our prior work suggested that short-acting drugs are more likely to show timing effects ^7^. This assessment relied on a binary yes/no measure of effect, ie. significance p< 0.05. We re-evaluated the impact of half-life via meta-regression ^22^ onto SMD, and found half-life to be a surprisingly poor predictor of effect size (slope = −0.0008, p = 0.8, R^2^ = 0) (Fig. 2C Supplement). Of note, our prior assessment included several studies of short-acting anti-cancer agents that were not part of this study because they reported only binary outcomes.

## Discussion

Treatment time effects can in theory arise from time-of-day influences on pharmacokinetics, pharmacodynamics, or underlying physiology. As genome-wide circadian datasets grow, so have predictions of medicine timing effects. In the past five years, ∼150 reviews mentioned ‘chronotherapy’ or ‘circadian medicine’ in the title or abstract, outpacing clinical evidence.

In this meta-analysis, we estimate a summarized effect of time of 0.41 standard deviations. What does this mean? While there is no straightforward way to assign reference values, an analysis of the most common drugs versus placebo found effects from SMD=0.1 to SMD=1.4 ^48^. In this context, timing effects from several studies were as large as many drug-versus-nothing effects. Unfortunately, the evidence for many of the interventions draws from a single trial of low precision. Which interventions are well-supported clinical opportunities that deserve focus?

Evidence from aspirin studies in this meta-analysis warrants future randomized controlled trials of aspirin timing in the prevention of cardiac events. Aspirin inhibits platelet reactivity and is widely used for cardiovascular disease prevention for this reason ^49^. Aspirin’s efficacy is attributed to its inhibition of cyclo-oxygenase-1 (COX-1) leading to inhibition of thromboxane A2 production by platelets ^50^. Interestingly, because the COX-1 inhibition is irreversible, aspirin’s therapeutic effect is traditionally thought to be sustained throughout the dosing interval once steady state is achieved. What, then, accounts for the dosing time effect on platelet reactivity observed in trials in this meta-analysis? One possibility is drug kinetics and platelet turnover. Aspirin has a short ∼1 h half-life and inhibits only the platelets present at the dosing time. New platelets are released at a rate of 10% per day ^51,52^ and it has been shown that the antiplatelet effect of aspirin declines during the 24 h dosing interval ^53,54^. Alternatively, the variable inhibition of other coagulation factors, such as prostaglandin I2, may modulate aspirin’s therapeutic platelet inhibition. Importantly, it is well established that platelet reactivity and coagulation peak in the morning ^41,42^. Approximately 300K recurrent cardiovascular events occur in the United States every year, with an excess of 40% during the morning hours ^55^. Even a small reduction in the morning peak could translate to a large decrease in recurrent events per year ^56^.

Studies of aspirin in platelet inhibition ^57^ underscore another point: the better time for treatment should be interpreted in the context of what was measured. For example, whereas morning dosing of proton pump inhibitors (PPIs) was more effective for *activity*-induced gastroesophageal reflux, evening dosing was more effective for *nocturnal* reflux ^58^. Constriction of the airway and pain also tend to peak at night. In this sense, optimal timing may be a matter of aligning drug exposure with a 24 h rhythmic symptom.

No area of drug timing has more published studies and debate than hypertension. Daily rhythms in blood pressure are well-documented. Disruption of these rhythms, in particular the loss of normal nighttime ‘dipping’, is associated with increased cardiovascular risk ^38–40^. Evidence that bedtime ingestion of antihypertensives is more effective for controlling nighttime blood pressure is therefore of wide interest. And interestingly, the greatest effects are seen with aspirin, findings that have been reproduced in mice ^59^. However, the clinical evidence draws largely from one source with effect sizes that are difficult to reconcile with the broader field. While confusing, we expect that continued advances in understanding the time-of-day-dependent mechanisms of cardiovascular function will clarify the role of time in treatment of heart disease.

## Limitations

SMD is an imperfect metric. Although insensitive to measurement units and thus ‘scale-free’, factors that influence the variance in a particular study can impact SMD and thereby compromise between-study comparisons. In addition, SMD is not robust to study outliers. Because outliers more strongly affect the variance than the mean, the presence of large outliers in a study can make SMD appear smaller than an analogous study without outliers. Alternative metrics that may better handle these issues require patient-level data, which was not available.

Where a study reported multiple outcomes, we selected a single one to use following an order of priority that favored statistical significance, as described in Methods. Our approach may therefore present a best-case scenario for timing effects. Alternative approaches that integrate *all* reported effects from a single study may be desirable but are challenging without patient-level data. Additionally, this meta-analysis did not consider binary outcomes, such as incidence of toxicity or survival, excluding ∼25 studies of chemotherapy timing in cancer. We collected data from these studies and provided it as a resource (Data file 1 in the Supplement).

## Conclusions

Despite many reports that attention to time-of-day can improve therapeutic outcomes, there is a critical need for both rigorous randomized controlled trials *and* analysis of real-world health data. Mechanistic insights from chronobiology will continue to drive hypotheses, but the burden of proof is clinical benefit. Published accounts of timing effects have tended to focus on *yes/no* significance. Future efforts to understand the range of likely effects should help clarify instances where consideration of time of day has the greatest medical potential.

## Data Availability

All data produced in this study are available upon request to the authors. However, all data produced can also be replicated by source code available on github and metadata available as a supplemental file.

## Supplement

**Fig 1 Supplement.**
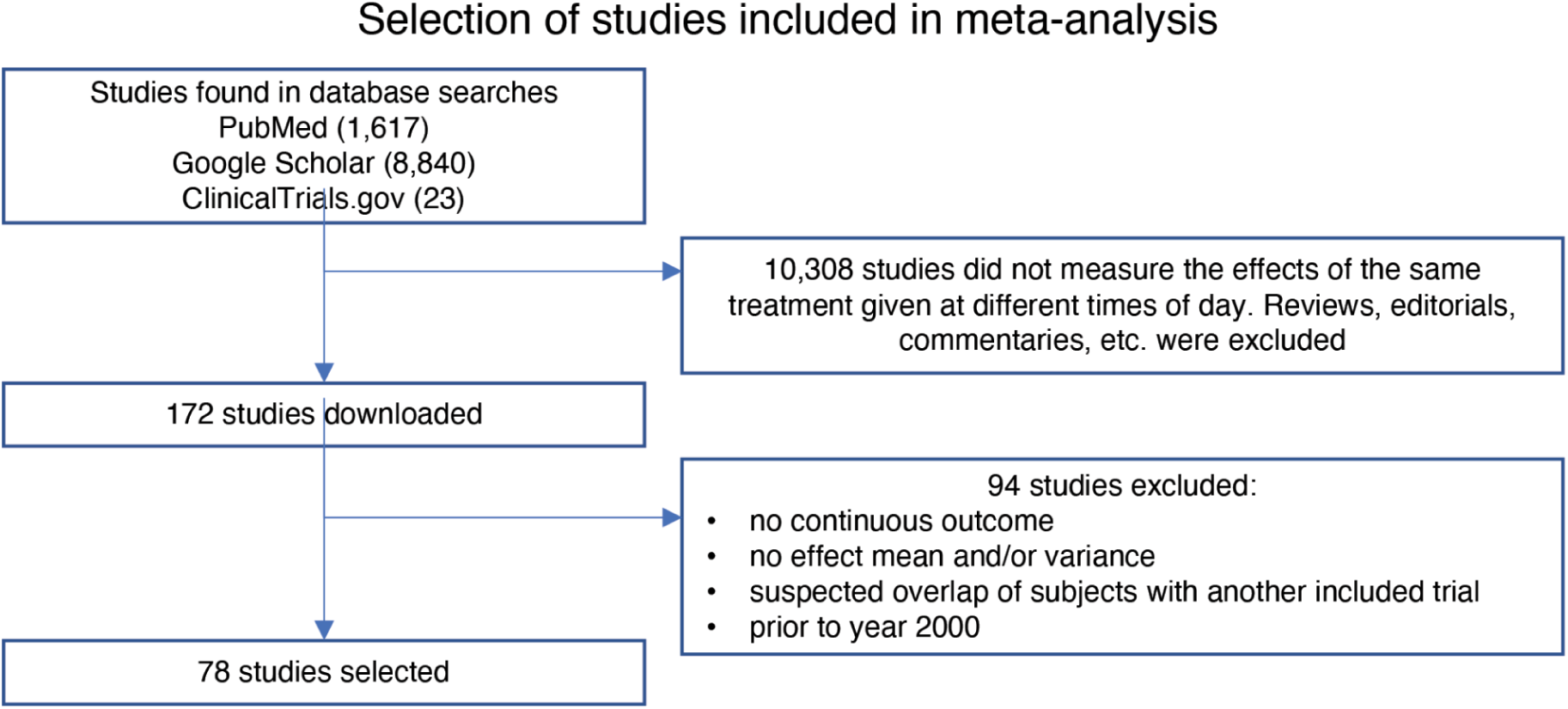
Selection of studies included in the meta-analysis (Methods).

**Fig 2 Supplement.**
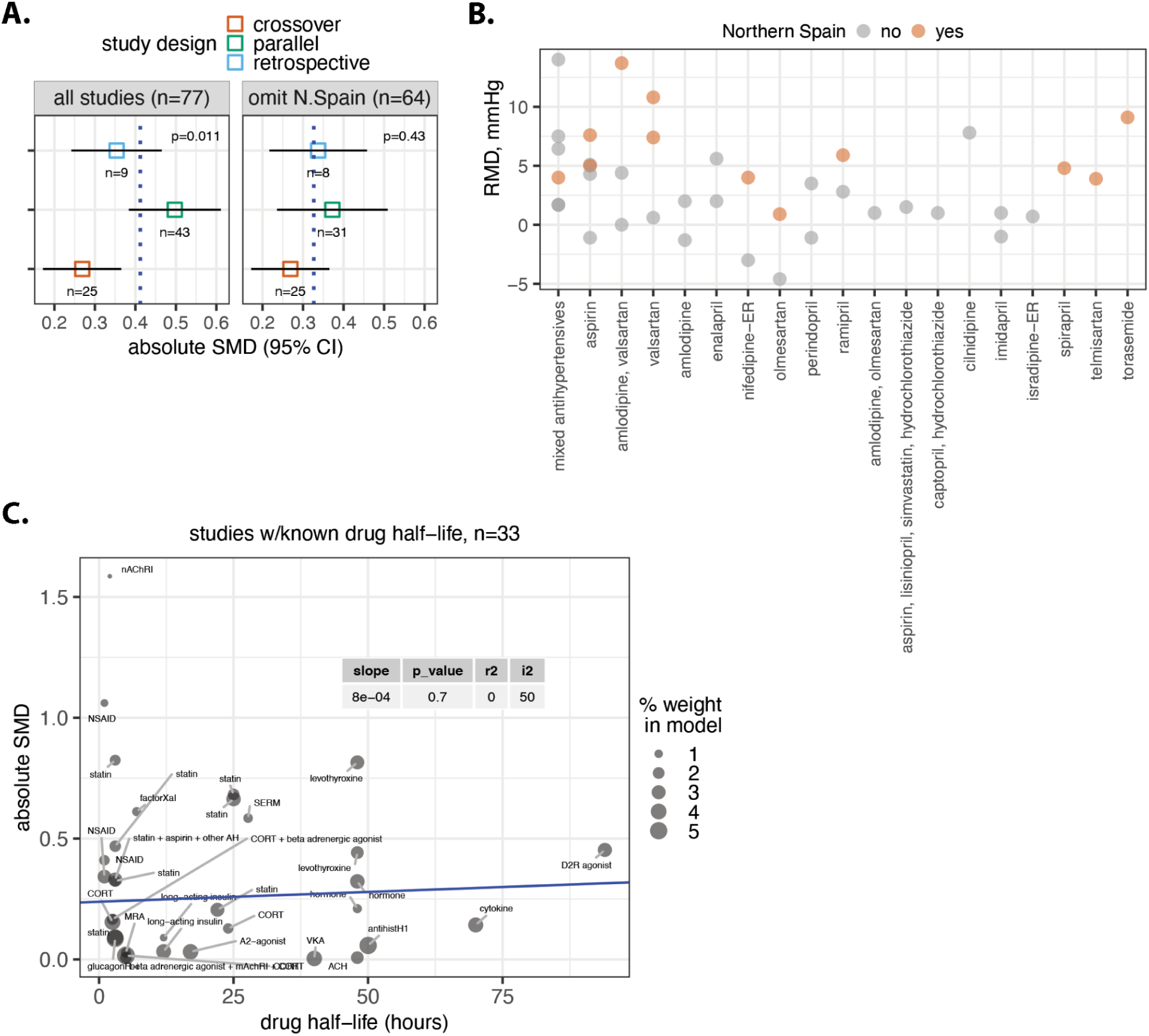
Influence of study features on timing effects. (**A**) Mixed-effects model pools the effect of each study design subgroup and compares between them. (**B**) Raw mean difference (RMD) for each distinct drug or drug combination that tested the effect of morning vs. evening dosing on nighttime systolic blood pressure. (**C**) Meta-regression of SMD onto drug half-life. *Point size*: study weight, ie., the contribution of each study to the estimated pooled effect. *Model coefficients*: R^2^ = percent of between-study variation explained by the model, I^2^ = residual heterogeneity, or the remaining variability between-studies not captured by half-life predictor, and p-value for difference between groups via Q-test. *Drug class abbreviations:* mAChRi: anticholinergic, A2-agonist: alpha 2 adrenergic receptor agonist, CORT: corticosteroid, GlucagonR-i: glucagon-like peptide-1 receptor agonist, VKA: vitamin K antagonist, SERM: selective estrogen receptor modulator; MRA: mineralocorticoid receptor antagonist.

## 78 Meta-analysis Studies

**Table 1 Supplement.** All studies included in this meta-analysis.

| OUTCOME AREA | REF | LINK |
| --- | --- | --- |
| Blood pressure | *Krasinska 2020* | 60 |
|  | *Hermida 2019* | 61 |
|  | *Zappe 2015* | 62 |
|  | *Ruben 2019* | 63 |
|  | *Rahman 2013* | 64 |
|  | *Tiwari 2020* | 65 |
|  | *Bonten 2015* | 56 |
|  | *Wang 2013* | 66 |
|  | *Ayala 2013* | 67 |
|  | *Zeng 2011* | 68 |
|  | *Kasiakogias 2015* | 69 |
|  | *Asmar 2011* | 70 |
|  | *Hermida 2010* | 71 |
|  | *Hoshino 2010* | 72 |
|  | *Hermida 2010* | 73 |
|  | *Hermida 2009* | 74 |
|  | *Hermida 2009* | 75 |
|  | *Hermida 2009* | 76 |
|  | *H. Mori 2013* | 77 |
|  | *Ushijima 2015* | 78 |
|  | *Hermida 2009* | 79 |
|  | *Hermida 2007* | 80 |
|  | *Hermida 2005* | 81 |
|  | *Hermida 2005* | 82 |
|  | *Qiu 2003* | 83 |
|  | *Serinel 2017* | 84 |
|  | *Hjortkjær 2016* | 85 |
|  | *Kitahara 2004* | 86 |
|  | *Hermida 2008* | 87 |
|  | *Hermida 2005* | 88 |
|  | *Kohno 2000* | 89 |
|  | *Farah 2013* | 90 |
|  | *Rossen 2014* | 91 |
|  | *Fujiwara 2017* | 92 |
|  | *Huangfu 2015* | 93 |
|  | *Kario 2016* | 94 |
|  | *Dimitrov 2012* | 95 |
|  | *Suomela 2015* | 96 |
|  | *Khodadoustan 2017* | 97 |
| Coagulation | *Krasinska 2018* | 98 |
|  | *Racca 2019* | 99 |
|  | *van Diemen 2021* | 57 |
|  | *Brunner-Ziegler 2016* | 100 |
|  | *Garrison 2020* | 101 |
|  | *NCT00515021* | NCT00515021 |
|  | *Ando 2013* | 102 |
| Lipid levels | *Chipchura 2018* | 103 |
|  | *Ozaydin 2006* | 104 |
|  | *Tharavanij 2010* | 105 |
|  | *Lafeber 2015* | 106 |
|  | *Wallace 2003* | 107 |
|  | *Lund TM 2002* | 108 |
|  | *Kim 2013* | 109 |
|  | *Yoon 2011* | 110 |
|  | *Plakogiannis 2005* | 111 |
|  | *Poulter 2018* | 112 |
| Thyroid function | *Bach-Huynh 2009* | 113 |
|  | *Bolk 2010* | 114 |
|  | *Elliott 2001* | 115 |
|  | *Rajput 2011* | (Rajput et al. 2011) |
| Pain | *Packard 2020* | 116 |
|  | *Buttgereit 2008* | 117 |
| Airway patency | *Calverley 2003* | 118 |
|  | *Zetterstrom 2008* | 119 |
|  | *NCT03108027* | NCT03108027 |
|  | *Kempsford 2016* | 120 |
|  | *Kempsford 2013* | 121 |
| Other | *Ren 2019* | 122 |
|  | *de Bree 2020* | 4 |
|  | *Montaigne 2017* | 123 |
|  | *Long 2016* | 3 |
|  | *Ahren 2013* | 124 |
|  | *Haye 2005* | 125 |
|  | *Patti 2020* | 126 |
|  | *Cheeseman 2007* | 127 |
|  | *Lucidi 2015* | 128 |
|  | *Garg 2004* | 129 |
|  | *Newcorn 2013* | 130 |

## Article Information

### Data deposition

Data and code for reproducing the analyses in this study have been deposited in GitHub (https://github.com/MarcDRuben/MetaAnalysis).

### Corresponding Authors

Marc D. Ruben PhD. Divisions of Human Genetics and Immunobiology, Department of Pediatrics, Cincinnati Children’s Hospital Medical Center, Cincinnati, USA.

John B. Hogenesch, PhD. Divisions of Human Genetics and Immunobiology, Department of Pediatrics, Cincinnati Children’s Hospital Medical Center, Cincinnati, USA.

### Author Contributions

Dr. Ruben had full access to all of the data in the study and takes responsibility for the integrity of the data and the accuracy of the data analysis.

*Concept and design*: Ruben

*Acquisition, analysis, or interpretation of data*: All authors

*Drafting of the manuscript*: Ruben, Francey, Wu, Hughey

*Critical revision of the manuscript for important intellectual content*: All authors

*Statistical analysis*: Ruben, Hughey, Wu

*Obtained funding*: Hogenesch

Supervision: Hogenesch, Hughey

